# Mutation in CDC42 gene set as a response biomarker for immune checkpoint inhibitor therapy

**DOI:** 10.1101/2023.11.10.23298355

**Authors:** Kun Wang, Yingying Zhang, Zhaoming Su, Bei Wang, Yuanyang Zhou, Xiaochu Tong, Chengying Xie, Xiaomin Luo, Sulin Zhang, Mingyue Zheng

## Abstract

**Background:** Immunotherapy has proven notably effective in treating tumors across diverse patient populations. However, some patients do not respond to immune checkpoint inhibitors (ICIs). Thus, there is a need for reliable biomarkers that can predict clinical responses to ICI treatment accurately.

**Methods:** Our focus is on CDC42, a protein that stimulates multiple signaling pathways, promoting tumor growth. We hypothesize that its defective function may indicate a patient’s response to ICI therapy. We consider CDC42, along with its downstream binding and effector proteins, as a gene set. This is because their mutation could result in defective CDC42 function. We investigated the mutations in the CDC42 gene set as a potential biomarker for clinical benefits from ICI treatment. We also examined whether the combined use of a CDC42 inhibitor and ICI could enhance the efficacy of ICI.

**Results:** The presence of mutations in the CDC42 gene set correlated with improved overall survival (OS: p = 2.9E-4) and progression-free survival (PFS: p = 2.92E-6). Furthermore, our analysis of immune response landscapes among different CDC42 gene set statuses supports its potential as a biomarker for ICI therapy. Animal experiments also revealed that combining the CDC42 inhibitor (ML141) with anti-PD-1 blockade can synergistically reduce tumor growth.

Conclusions

Our study suggests that the CDC42 gene set could serve as a novel biomarker for the clinical response to ICI treatment. This finding also provides insight into the potential of combining ICI and CDC42 inhibitor use.

## Background

Immune inhibitor therapy, such as anti-programmed cell death (ligand) 1 [PD-(L)1] and anti-cytotoxic T lymphocyte antigen 4 (CTLA-4) drugs, has been successful in cancer therapy and improving long-term survival for patients. However, the effectiveness of immune checkpoint inhibitor (ICI) drugs can vary greatly among different patients due to tumor heterogeneity. While these drugs have shown positive effects in some patients, 60%-80% of patients do not respond clinically to them[1]. Therefore, it is important to identify predictive biomarkers that can indicate patients’ clinical benefit[2].

CDC42 is a type of ras homologous (rho) GTPase. Previous studies have reported that CDC42 simulates tumor genesis, progression, invasion and metastatic[3]. In a previous study by Kalim et al., it was reported that inhibiting CDC42 activity in regulatory T cells (Tregs) can enhance anti-tumor immunity[4]. While Kalim et al. report that the immuno-effect of the CD42 inhibitor outweighs any tumor cell-intrinsic effect[4], it has also been reported that a low level CDC42 in the serum can predict the clinical response to ICI in patients with advanced hepatocellular carcinoma (HCC) [5] and advanced cervical cancer[6]. Therefore, it is valuable to explore the defective function of CDC42 signaling in tumors beyond Tregs and whether it can raise the probability of a response to ICI. If the defective function of CDC42 signaling is a biomarker for ICI therapy, it could provide further insight into the combined use of ICI and CDC42 inhibitor.

The function of CDC42 in control cell growth and polarity not only depends on itself but also on its binding protein and effector protein. Therefore, we consider CDC42, its binding protein, and effector protein as a gene set and investigate their potential as a biomarker for indicating the clinical benefit of ICI therapy, i.e., explore the defectiveness of CDC42 function’s biomarker role indirectly. We examine whether there is a significant difference in overall survival (OS) and progression-free survival (PFS) among patients with different CDC42 gene set statuses. And we do bootstrap based on collected ICI therapy datasets. Using the ICI therapy datasets and their bootstrap samples, we assess whether the CDC42 gene set status can differentiate the clinical benefit of ICI treatment in patients. To further explain the predictive performance of the CDC42 gene set status, we examine the intrinsic and extrinsic immune response landscapes among its status. Additionally, we investigate whether the CDC42 gene set status significantly differentiates signature levels reported to influence the efficacy of ICI therapy, such as the level of CD8 T cell infiltration.

Furthermore, to validate whether defective CDC42 function can serve as a biomarker for ICI, we investigated whether ML141 could increase the survival time of mice. We selected mice with 4T1 breast carcinoma, which has been shown to be highly resistant to anti-PD-1 or anti-CTLA-4 therapy for the experiment[7]. ML141 is a selective and non-competitive inhibitor of CDC42[8], which was utilized to simulate the defective function of CDC42. Our study also aims to demonstrate the potential of CDC42 inhibitors in improving the anti-tumor effects of ICI, particularly in cases where tumors exhibit resistance to ICI therapy. Overall, our study explores the role of CDC42 gene set status as a biomarker for ICI therapy and seeks evidence to support the use of a CDC42 inhibitor to enhance the efficacy of the ICI inhibitors.

## Methods

### Materials

We collected nine whole exome sequencing (WES) data for biomarker discovery. The Miao2019 cohort consists of renal clear cell carcinoma patients treated with anti-PD-1 drugs[9]. The Hugo and Riaz cohorts comprise melanoma patients treated with anti-PD-1 drugs [10, 11]. The Miao2018 cohort consists of pan-cancer patients treated with either 1) anti-cytotoxic T lymphocyte-associated protein-4 (CTLA-4) drugs, 2) anti-PD-1 drugs, or 3) a combination of both anti-CTLA-4 and anti-PD-1 drugs [12]. The Rizvi cohort comprises non-small cell lung cancer (NSCLC) patients treated with anti-PD-1 drugs[13]. The Snyder and Van Allen cohorts consist of melanoma patients treated with anti-CTLA-4 drugs [14, 15]. The Hellmann cohort comprises non-small cell lung cancer patients treated with both anti-CTLA-4 and anti-PD-1 drugs[16]. The Liu cohort comprises melanoma patients treated with anti-PD-1 drugs[17](see supplementary material Table S1). We downloaded eight WES datasets and corresponding clinical information from the cBioPortal database (https://www.cbioportal.org). The Riaz cohort was obtained from the original literature[12].

The collected data pertains to the drug response of cancer patients undergoing ICI therapy. In order to combine this data from multiple sources, we utilized the processing method described by Zhang et al[18]. Initially, we excluded three tumor types with a sample size of less than 10. We also removed 33 samples that had a non-evaluable response (NE), 7 samples that were not profiled and 7 samples classified as “OTHER CONCURRENT THERAPY”. Furthermore, we eliminated 151 duplicate samples in the Miao2018 cohort. This cohort had 27 overlapping samples with the Rizvi cohort, 37 with the Snyder cohort, and 87 with the Van Allen cohort.

Table 1 summarizes the characteristics of the filtered data. The ICI therapy dataset includes five tumor types: Melanoma (n=422), Non-Small Cell Lung Cancer (128), Renal Cell Carcinoma (35), Bladder Cancer (27) and Head and Neck Cancer (10). The ICI therapy dataset includes the following types of drug treatment: anti-PD-1 (306), anti-CTLA-4 (174) and anti-CTLA-4 + anti-PD-1 (142). The proportion of CDC42 gene set mutation in the ICI therapy dataset is 18%.

**Table 1.**
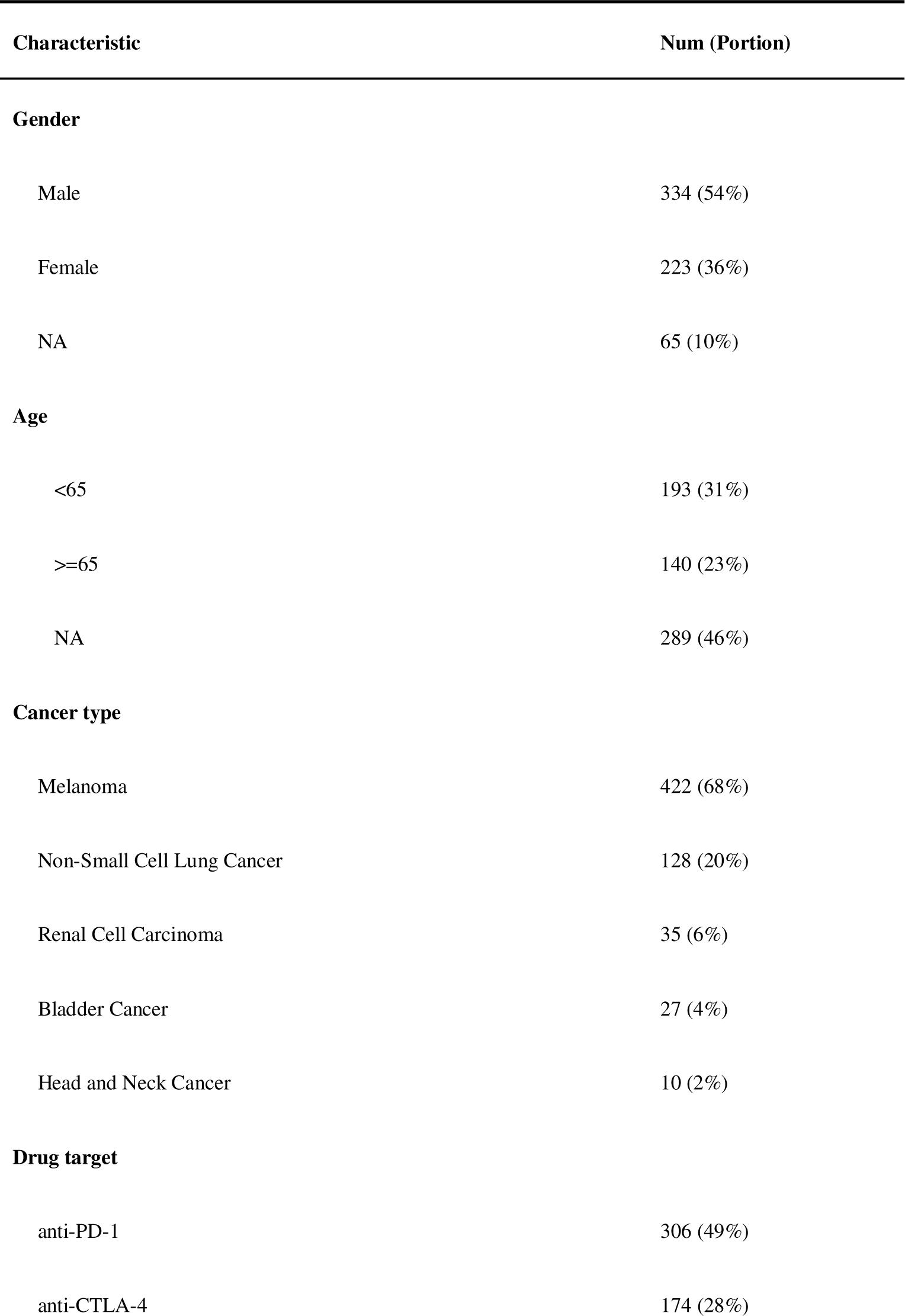
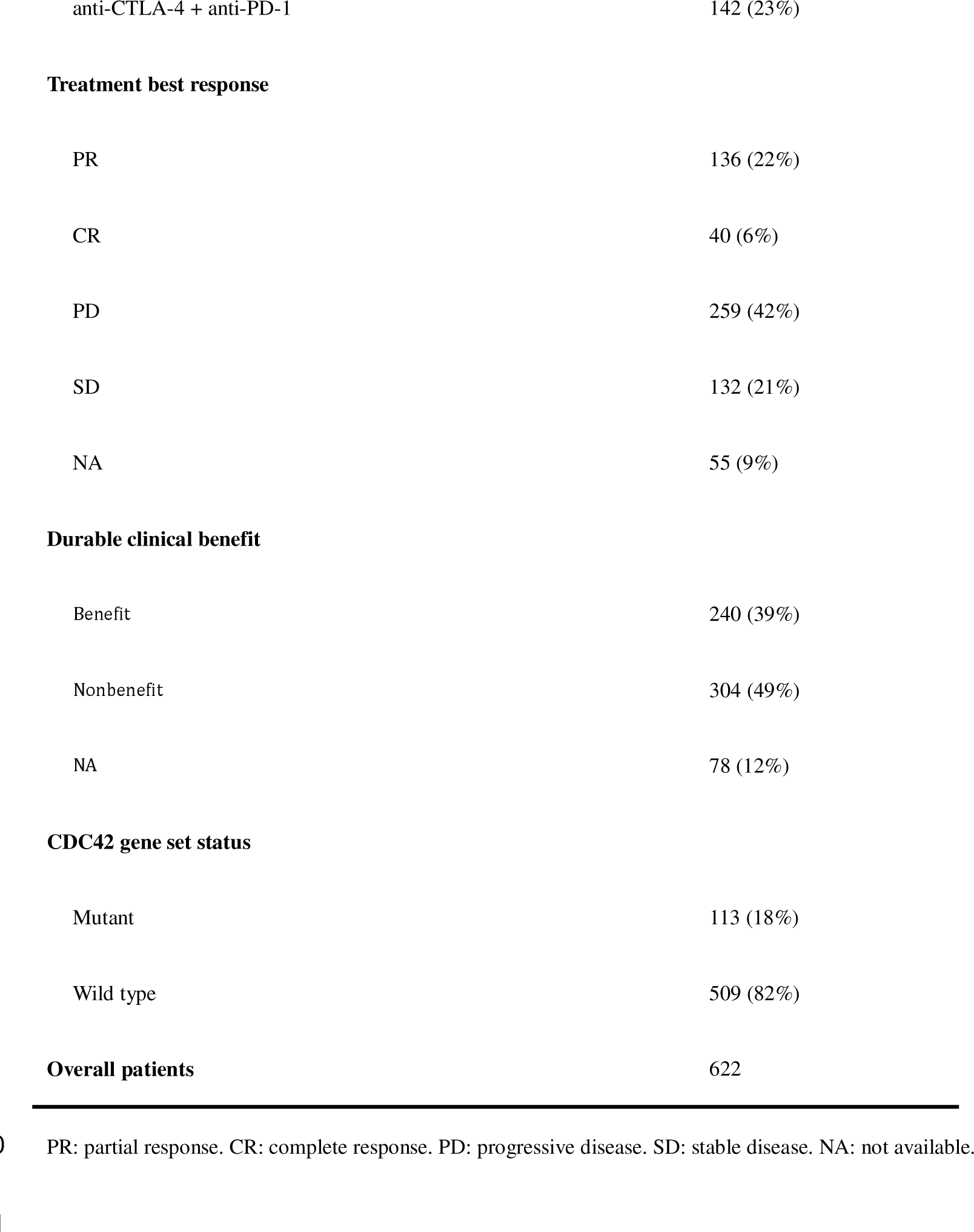
Characteristics of the ICI therapy dataset.

| Characteristic | Num (Portion) |
| --- | --- |
| **Gender** |  |
| Male | 334 (54%) |
| Female | 223 (36%) |
| NA | 65 (10%) |
| **Age** |  |
| <65 | 193 (31%) |
| >=65 | 140 (23%) |
| NA | 289 (46%) |
| **Cancer type** |  |
| Melanoma | 422 (68%) |
| Non-Small Cell Lung Cancer | 128 (20%) |
| Renal Cell Carcinoma | 35 (6%) |
| Bladder Cancer | 27 (4%) |
| Head and Neck Cancer | 10 (2%) |
| **Drug target** |  |
| anti-PD-1 | 306 (49%) |
| anti-CTLA-4 | 174 (28%) |
| anti-CTLA-4 + anti-PD-1 | 142 (23%) |
| **Treatment best response** |  |
| PR | 136 (22%) |
| CR | 40 (6%) |
| PD | 259 (42%) |
| SD | 132 (21%) |
| NA | 55 (9%) |
| **Durable clinical benefit** |  |
| Benefit | 240 (39%) |
| Nonbenefit | 304 (49%) |
| NA | 78 (12%) |
| **CDC42 gene set status** |  |
| Mutant | 113 (18%) |
| Wild type | 509 (82%) |
| **Overall patients** | **622** |

PR: partial response. CR: complete response. PD: progressive disease. SD: stable disease. NA: not available.

Furthermore, we collected data from 32 types of solid cancer data from The Cancer Genome Atlas (TCGA) to conduct further analysis on the CDC42 gene set status as a biomarker. This data includes WES data, RNA-seq data, and patients’ overall survival time. The WES and RNA-seq data were obtained using TCGAbiolinks[19], and the survival time data were collected from Liu et al[20]. Additionally, we obtained Cibersort immune infiltration values and TCR Shannon for each TCGA cancer sample from Thorsson et al[21].

### CDC42 gene set mutation definition

In this study, we defined the CDC42 gene set as a set of genes that includes CDC42, CDC42 binding protein kinase alpha (CDC42BPA), CDC42 binding protein kinase beta (CDC42BPB), CDC42BPG CDC42 binding protein kinase gamma (CDC42BPG), CDC42 effector protein 1 (CDC42EP1), CDC42 effector protein 2 (CDC42EP2), CDC42 effector protein 3 (CDC42EP3), CDC42 effector protein 4 (CDC42EP4), CDC42 effector protein 5 (CDC42EP5), CDC42 small effector 1 (CDC42SE1), CDC42 small effector 2 (CDC42SE2). These genes are involved in CDC42 function, and their mutation may affect CDC42’s signal. If any gene in this gene set undergoes a non-synonymous mutation, the CDC42 gene set status is defined as mutated, which means the defective function of CDC42 to some extent.

### Clinical endpoint analysis

The objective response rate (ORR) was defined as the proportion of patients who received ICI therapy and achieved a complete response (CR) or partial response (PR)[22]. Durable clinical benefit (DCB) was defined as a CR, PR, or stable diseases (SD) that lasted for more than 6 months[23].

### Immune cell fraction analysis

We obtained the leukocyte fraction from Thorsson et al[21]. The lymphocyte fractions were aggregated by using the cibersort estimate, including B cells naïve, B cells memory, T cells CD4 naïve, T cells CD4 memory resting, T cells CD4 memory activated, T cells follicular helper, Tregs, T cells gamma delta, T cells CD8, NK cells resting, NK cells activated, and Plasma cells[21]. The molecular estimate for tumor-infiltrating lymphocyte (TIL) fraction was obtained by multiplying the aggregated lymphocyte fraction from the cibersort estimate with the leukocyte fraction obtained from Thorsson et al. The estimate for TIL fraction images was obtained from Saltz et al[24].

### Immune signatures analysis

We obtained 29 immune signatures from He et al[25] and performed single-sample gene set enrichment analysis (ssGSEA) using the “GSVA” R package[26] based on these signatures.

### GSEA analysis

We used TCGA RNA-seq data and the DESeq2 package[27] to identify differentially expressed genes.

Subsequently, we conducted GSEA analysis on the Kyoto Encyclopedia of Genes and Genomes (KEGG) pathway using the clusterProfiler package[28].

### Estimation of cytolytic activity

We estimated cytolytic activity (CYT) based on the method described by Rooney et al[29]. This involves calculating the geometric mean of granzyme A (GZMA) and perforin 1 (PRF1) expression.

### Mutation and neoantigens analysis

For this study, we assessed tumor mutational burden (TMB) using the number of non-synonymous mutations. The data for nonsilent mutation, silent mutation, single nucleotide variation (SNV) neoantigens and indel neoantigens were obtained from Thorsson et al[21].

### Statistical analysis

We used the two-sided Fisher’s exact test to explore the difference between CDC42 gene set status and clinical benefit. To further assess the potential of CDC42 gene set as a biomarker for predicting the clinical benefits of ICI, we employed bootstrapping to generate 1000 bootstrap samples for both CDC42 gene set mutation and wild type patients. This allowed us to obtain empirical distributions for ORR and DCB[30]. Subsequently, we then compared the 95% confidence intervals of ORR and DCB based on the CDC42 gene set status. The two-sided Wilcoxon rank sum test was used to compare the TMB and neoantigen load (NAL) of ICI therapy data. We also used the two-sided Wilcoxon rank sum test to compare TCGA gene expression levels, mutation rate, neoantigens, cell fraction, immune signatures, TCR Shannon and CYT between the CDC42 gene set mutation group and the CDC42 gene set wild type group. Additionally, we plotted the KM curve of PFS and OS using the logrank test based on CDC42 gene set status, which used the `X`^2^ test statistic to calculate P values[31]. Fisher’s exact test was implemented using the python package scipy[32]. The logrank test, Cox Proportional-Hazards analysis, and Wilcoxon rank sum test were implemented using the survminer package and ggsignif package in R version 4.2.3 (https://www.r-project.org).

### Animal experiment

All procedures performed on animals were conducted in accordance with the Institutional Animal Care and Use Committee at the Shanghai Institute of Materia Medica, Chinese Academy of Sciences (IACUC Issue NO. 2023-10-ZMY-03). For the pharmacodynamics experiment, BALB/c mice (6-8 weeks old) were purchased and inoculated subcutaneously with 1×10^6^ 4T1 tumor cells into the right side of the mice’s axilla. The animals were divided into four groups irregularly once the tumor volume reached approximately to 100 mm^3^. ML141 (#HY-12755, MedChemExpress) was administered in a solution containing PEG300, dimethyl sulfoxide, and PBS [40/5/55 (v/v/v)]. The mice were then treated intraperitoneally with or without ML141 (30 mg/kg once a day) and/or 150 _μ_g/mouse of anti-PD-1 antibody (#-BE0273, Bio X Cell) every other day for one injection. Tumor volumes were calculated using the formula: V= (length×width^2^)/2. Body weights and tumor volumes of the mice were measured daily. The tumor growth inhibition (TGI) value was calculated using the formula: TGI= [1-Relative Tumor Volume (Treatment)/Relative Tumor Volume (Vehicle)] ×100%.

### Flow cytometry analysis

The tumor tissues were firstly digested into single cells using a digestion solution containing 0.001% hyaluronidase, 0.1% collagenase, 0.002% DNase, 120 _μ_M MgCl2, and 120 _μ_M CaCl2 in RPMI 1640 medium. Subsequently, red blood cells were lysed using ammonium chloride for 3 minutes and then the cell samples were stained with Fixable Viability Stain 700 (#564997, BD). The Fc receptors were blocked with TruStain FcX™ (anti-mouse CD16/32) antibody (#101320, Biolegend) and stained with the following antibodies: APC-Cy7 rat anti-mouse CD45 (#557659, BD), FITC CD3 monoclonal antibody (17A2) (#11-0032-82, Invitrogen), and Brilliant Violet 421 anti-mouse CD8a antibody (#100738, Biolegend). The stained cells were analyzed using the Agilent Novocyte 3000 instrument, and all data were analyzed with FlowJo software.

## Results

### Mutation in CDC42 gene set was associated with improved clinical outcomes for ICI therapy

As shown in Fig. 1 a, the ORR of patients in the CDC42 gene set mutation group (ORR = 53%, 53/100) was significantly higher (p = 6.59E-7) compared to the CDC42 gene set wild type group (ORR = 26.34%, 123/467). Additionally, the DCB of patients in the CDC42 gene set mutation group (DCB = 64.21%, 61/95) was also significantly higher (p = 2.12E-5) than the wild type group (DCB = 39.87%, 179/449). Moreover, CDC42 gene set mutation patients had significantly longer overall survival time (p = 2.9E-4, HR = 0.52, 95%CI = 0.36-0.75) and progression-free survival time (p = 2.92E-6, HR = 0.46, 95%CI = 0.33-0.64) compared to the CDC42 gene set wild type group.

**Fig. 1.**
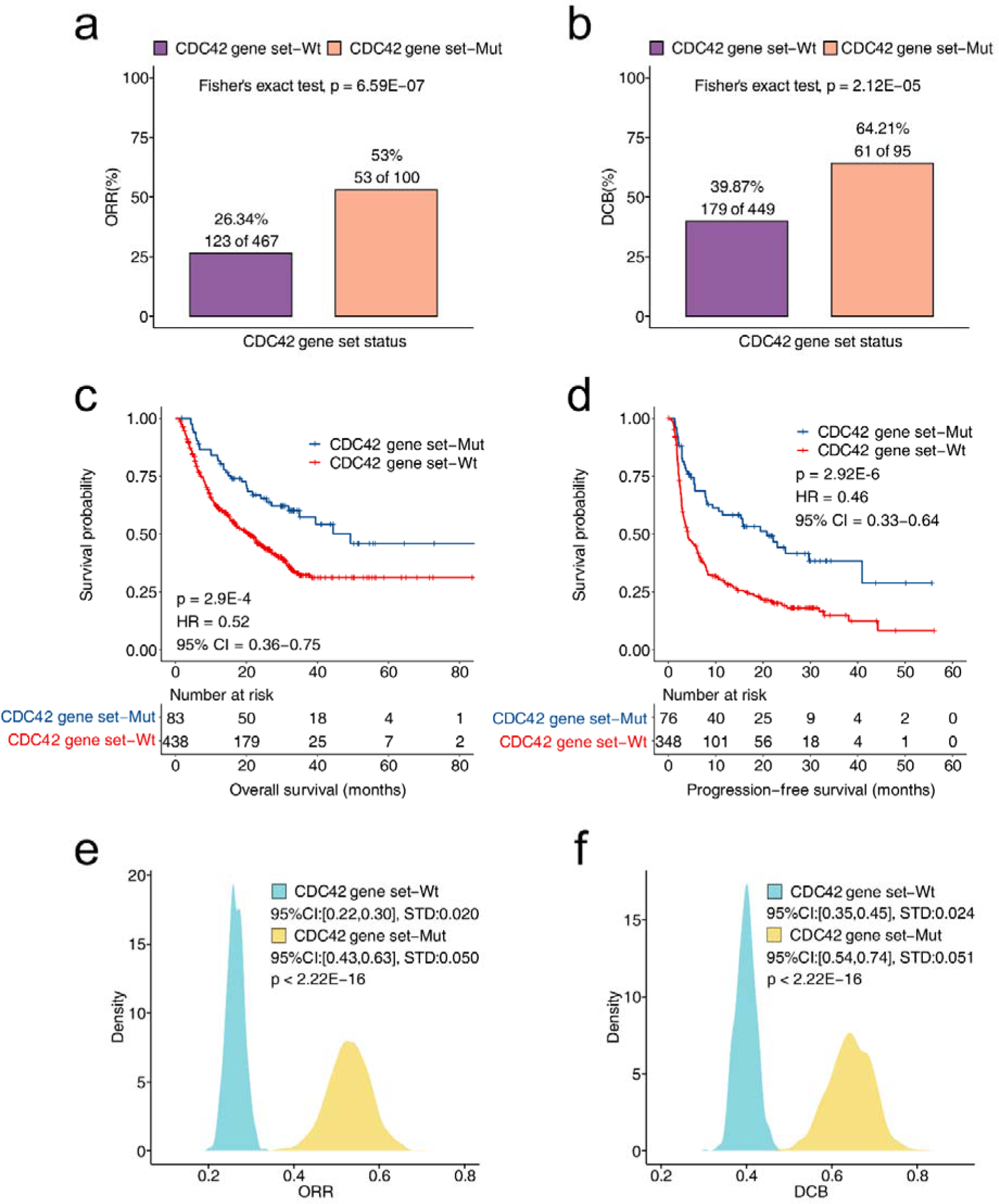
Analysis of CDC42 gene set mutation as a biomarker for ICI therapy. **a**, The differences in ORR between CDC42 gene set mutation and CDC42 gene set wild type groups. **b**, The differences in DCB between CDC42 gene set mutation and CDC42 gene set wild type groups. **c**, The KM curve of OS based on the CDC42 gene set status. **d**, The KM curve of PFS based on the CDC42 gene set status. **e**, The distribution of mean values of ORR based on bootstrap samples. **f,** The distribution of mean values of DCB based bootstrap samples. ORR, objective response ratio; DCB, durable clinical benefit; KM, Kaplan-Meier; OS, overall survival; PFS, progression-free survival.

We divided the collected ICI therapy dataset into two groups: CDC42 gene set mutation group and CDC42 gene set wild type group. For each patient group, we sampled 1000 times to further explore ORR’s difference in different CDC42 gene set statuses. The same operation was also performed on DCB. As shown in Fig. 1e, the mean values distribution of ORR was significantly different between the two patient groups (p < 2.22E-16). The 95% confidence interval (CI) was 0.22-0.30, with a standard deviation (STD) was 0.02 in CDC42 gene set wild type group. In CDC42 gene set mutation group, the 95%CI was 0.43-0.63, and the STD was 0.05. Similarly, in Fig. 1f, the mean values distribution of DCB was significantly different between the two patient groups (p < 2.22E-16). The 95%CI was 0.35-0.45, with a STD of 0.024 in CDC42 gene set wild type group. In CDC42 gene set mutation group, the 95%CI was 0.54-0.74, and the STD was 0.051. Based on these results, it can be inferred that patients with CDC42 gene set mutations are more likely to have clinical benefits from receiving ICI treatment. Overall, the results in Fig. 1 suggest that the CDC42 gene set mutation could serve as a biomarker for the clinical response of ICI treatment.

### Assessment of intrinsic immune response landscapes in CDC42 gene set wild type and mutation tumors

We initially examined the relationship between CDC42 gene set status and immunogenicity in the ICI therapy cohort. As shown in Fig. 1a and Fig. 1b, the levels of TMB and NAL in the CDC42 gene set mutation group were significantly higher than those in the CDC42 gene set wild type group (TMB: p < 2.22E-16, NAL: p = 8.1E-14). These findings indicate a strong association between CDC42 gene set mutations and increased immunogenicity, as well as a higher likelihood of positive responses to ICI therapy. We also explored the relationship between CDC42 gene set status and immunogenicity in the TCGA cohort. Compared to CDC42 gene set wild type tumors, both the nonsilent mutation rate and the silent mutation rate were significantly higher than in CDC42 gene set mutation tumors (p < 2.22E-16, Fig. 2c, d). Additionally, both SNV neoantigens and indel neoantigens were significantly more abundant in in CDC42 gene set mutation tumors compared to CDC42 gene set wild type tumors (p < 2.22E-16, Fig. 2e, f). These results in the TCGA cohort further support the notion that the CDC42 gene set mutations are associated with enhanced tumor immunogenicity.

**Fig. 2.**
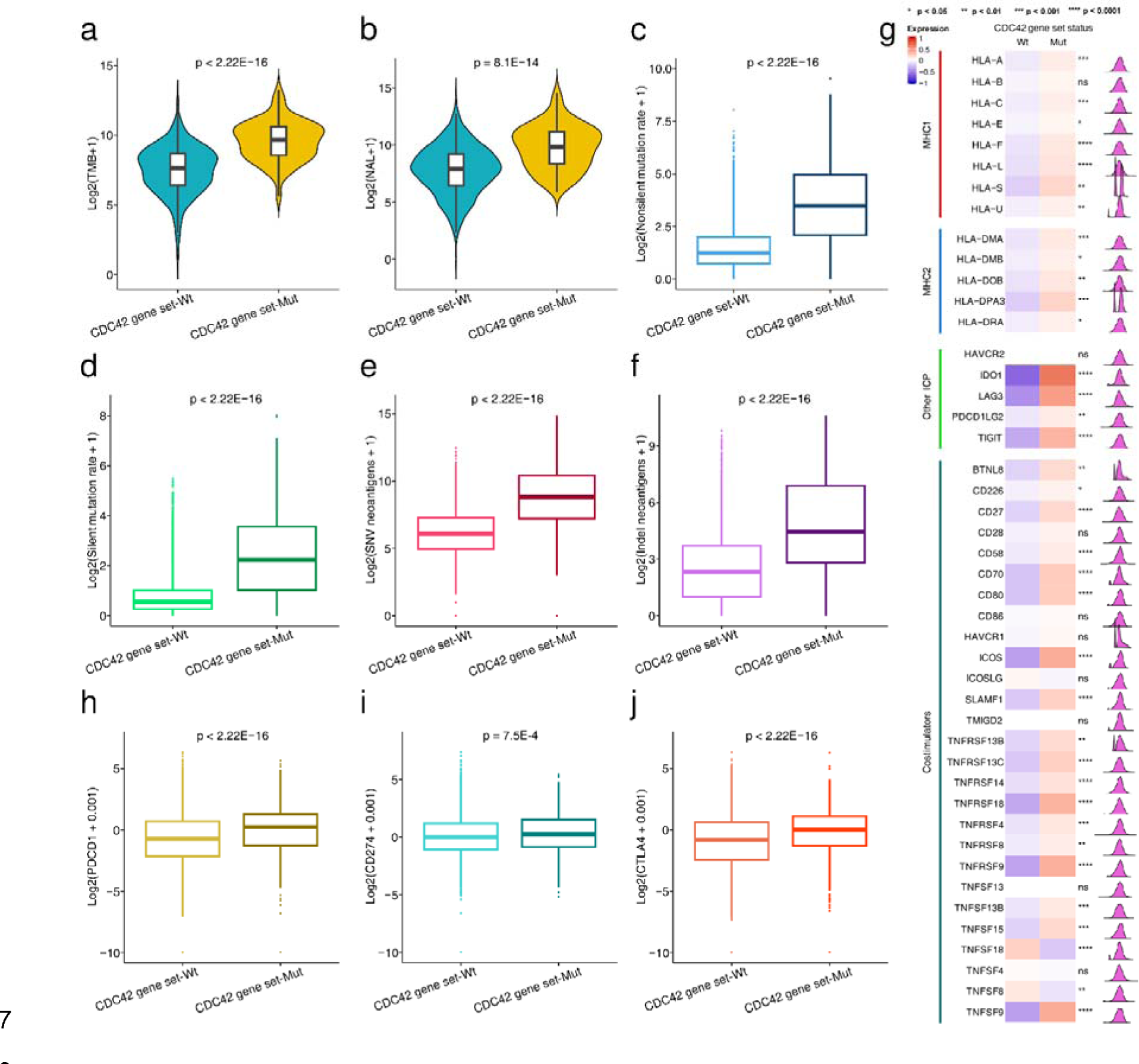
Analysis of intrinsic immune response landscapes in CDC42 gene set wild type and mutation tumors. Comparison of **a.** TMB, **b.** NAL between CDC42 gene set wild type and CDC42 gene set mutation groups in the ICI therapy cohort. Comparison of **c.** nonsilent mutation rate, **d.** silent mutation rate, **e.** SNV neoantigens, **f.** indel neoantigens, **g.** expression of MHC and other ICP molecules and costimulators, **h.** expression of PDCD1, **i.** expression of CD274, **j.** expression of CTLA-4 between CDC42 gene set wild type and CDC42 gene set mutation groups in the TCGA cohort. TMB, tumor mutation load; ICI, immune checkpoint inhibitor; NAL, neoantigen load; SNV, single nucleotide variant; ICP, immune checkpoint. *P < 0.05, **P < 0.01, ***P < 0.001, ****P < 0.0001

Then, we investigated the relationship between CDC42 gene set status and the expression of immune-related molecules, including two class MHC molecules, immune checkpoint, and co-stimulators. We found that immune checkpoint genes PDCD1, CD274 and CTLA-4 were upregulated in the CDC42 gene set mutation group. as shown in Fig 2h-j. Additionally, we observed significantly higher expression of MHC1, MHC2, other immune checkpoints (ICPs), and co-stimulators in CDC42 gene set mutation tumors compared to CDC42 gene set wild type, as shown in Fig. 2g. Previous studies have suggested that higher expression of immune checkpoint-related genes is indicative of a better response to ICIs therapy[33, 34]. MHC1 plays a crucial role in presenting antigens to CD8 T cells, and its down-regulation is associated with resistance to ICIs[35]. Moreover, positive expression of MHC2 correlates with a response to ICIs therapy[36]. Co-stimulators can promote T cell activation and survival, and activation of co-stimulatory pathways enhances checkpoint inhibition[37, 38]. In summary, these results demonstrate that CDC42 gene set mutation is a strong predictive biomarker for ICI therapy response.

### Assessment extrinsic immune response landscapes in CDC42 gene set wild type and mutation tumors

The different situations of immune cell infiltration result in different clinical outcomes of ICI therapy[39]. Therefore, we investigated the difference in tumor microenvironment (TME) between CDC42 gene set wild type and mutation tumors. This included analyzing immune cell score, signatures representing their function, and differential gene expression related to immune cell and ICI therapy efficiency.

As shown in Fig. 3a and 3b, the leukocyte fraction and lymphocyte fraction in CDC42 gene set mutation tumors were significantly higher than those in CDC42 wild type gene set tumors (leukocyte fraction, p = 5.7E-06; lymphocyte fraction, p = 6.3E-06). As TIL is crucial for killing tumors[40], TIL fractions estimated at both molecular and image levels were compared. Fig. 3c shows that TIL fractions (molecular estimate) in CDC42 gene set mutation tumors are significantly higher than those in CDC42 gene set wild type tumors (p = 9.5E-3). Fig. 3d shows that TIL fractions (images estimate) in CDC42 gene set mutation tumors are significantly higher than in those in CDC42 gene set wild type tumors (p = 3E-06). These results indicate that CDC42 gene set mutation tumors are more likely to be recognized and killed by immune cells than CDC42 gene set wild type tumors.

**Fig. 3.**
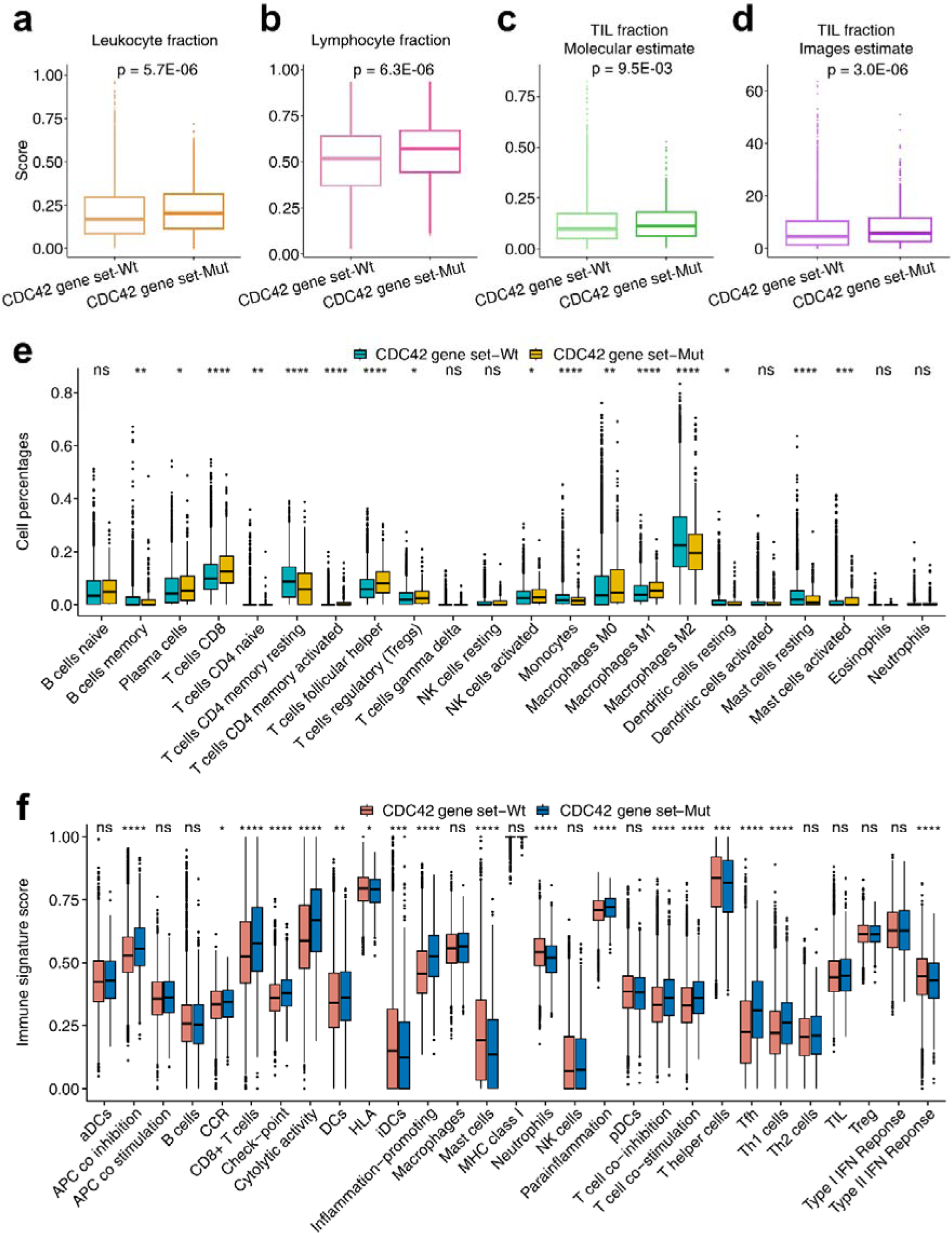
Analysis of extrinsic immune response landscapes of CDC42 gene set wild type and CDC42 gene set mutation tumors in the TCGA cohort. Comparison of **a.** leukocyte fraction, **b.** lymphocyte fraction, **c.** TIL fraction based on molecular estimates, **d.** TIL fraction based on images estimates, **e.** immune cell infiltration, **f.** 29 immune signatures estimated through the ssGSEA method between CDC42 gene set wild type and CDC42 gene set mutation tumors. *P < 0.05, **P < 0.01, ***P < 0.001, ****P < 0.0001

Cibersort scores based on the TCGA cohort were also compared between CDC42 gene set wild type and mutation tumors. As shown in Fig. 3e, the percentages of immune cell types were compared in detail. We found significant differences in most of the immune cell scores between CDC42 gene set wild type and mutation tumors. For example, the CD8 T cell and macrophage M1 cell scores in the CDC42 gene set mutation type were significantly higher than that in the CDC42 gene set wild type tumors. These findings are consistent with previous reports that CD8 T cell are key determinants of response to ICI, and macrophage M1 cells are related to T cell stimulation and ICI therapy[34, 41]. Fig. 3f shows ssGSEA results based on 29 immune signatures. We found that CD8 T cell and checkpoint signatures in CDC42 gene set mutation type tumors were significantly higher than those in CDC42 gene set wild type tumors. These results are also consistent with previous reports that CD8 T cells are key determinants of response to ICI and higher expression of immune checkpoint-related genes is more likely to benefit clinically from ICIs treatment[33]. Fig. 4a further shows that immune signatures were significantly enriched in CDC42 gene set mutation tumors compared to CDC42 gene set wild type tumors.

**Fig. 4.**
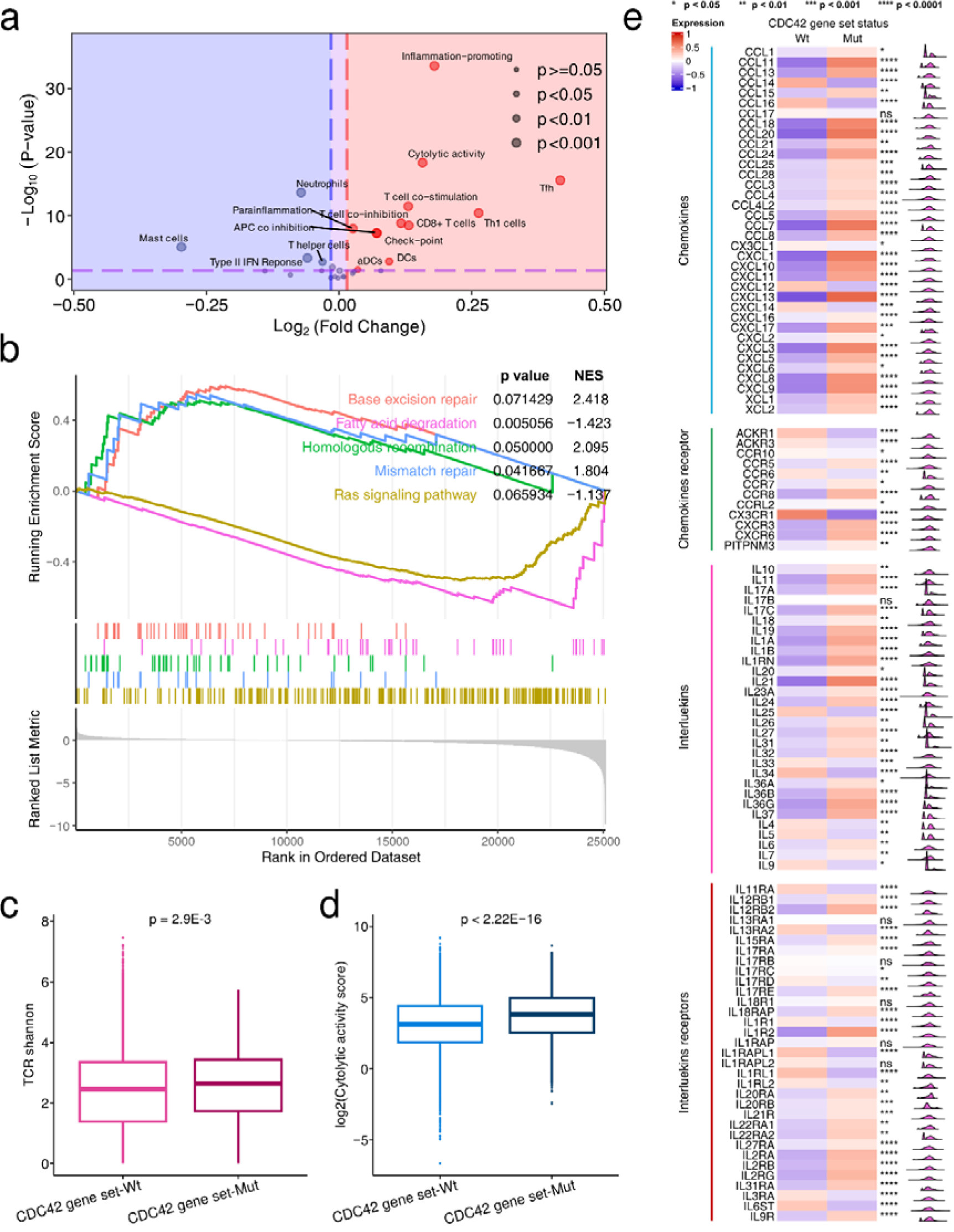
Analysis of gene expression related to immune cell between CDC42 gene set wild type and CDC42 gene set mutation tumors in the TCGA cohort. **a**, Volcano plots showing the analysis of 29 immune signatures estimated by the ssGSEA method for CDC42 gene set wild type and CDC42 gene set mutation tumors. **b**, GSEA analysis results based on CDC42 gene set status, with gene sets having an FDR (Benjamini-Hochberg method) lower than 0.25 considered significantly enriched. **c**, Comparison of the TCR Shannon score between CDC42 gene set wild type and CDC42 gene set mutation tumors. **d**, Comparison of the cytolytic activity score between CDC42 gene set wild type and CDC42 gene set mutation tumors. **e**, Comparison of the chemokines (and receptors) and interleukins (and receptors) between CDC42 gene set wild type and CDC42 gene set mutation tumors. *P < 0.05, **P < 0.01, ***P < 0.001, ****P < 0.0001.

We further conducted GSEA analysis, as well as expression analysis of chemokines and chemokines receptors, interleukins and interleukins receptors, TCR, and cytolytic activity score based on CDC42 gene set status. In Fig. 4b, we observed enrichment of base excision repair, homologous recombination, mismatch repair pathway enriched in CDC42 gene set mutation tumors, while fatty acid degradation and Ras signaling pathway were enriched in CDC42 gene set wild type tumors. These results align well with previous reports. For example, Jiang et al reported that DDR pathways are associated with the response to ICIs treatment[42]. Ward et al reported that activation of the Ras signaling pathway leads to an immunosuppressive tumor microenvironment, hindering T cells activation and infiltration, thus affecting the therapeutic efficacy of ICI[43]. Li et al reported that increased lipid content is correlated with a favorable ICI response[44], suggesting that higher lipid accumulation may indicate a higher likelihood of a positive response to ICI therapy. In Fig. 4c, we found a higher TCR Shannon score in CDC42 gene set mutation tumors compared to that in CDC42 gene set wild tumors (p = 2.9E-3). Higher TCR diversity may indicate that T cells can recognize more neoantigens, and studies have shown that patients with higher TCR diversity scores have more favorable clinical responses to ICI treatment[45]. The cytolytic activity score in CDC42 gene set mutation patients was significantly higher than in CDC42 gene set wild type patients (Fig.4d, p <2.22E-16). CYT is upregulated during T cell activation[46], indicating that upregulated CYT leads to more effective tumor killing. As shown in Fig. 5e, most chemokines in CDC42 gene set mutation patients were significantly higher than in CDC42 gene set wild type patients. Previous studies have reported that CXCL9, CXCL10 and CXCL11 can enhance T cell infiltration, thereby improving the therapeutic efficacy of ICI interventions[47–49]. The expression of CXCL13 can generate effector T cells and is closely associated with the response to ICIs treatment[50]. There were also significant differences in interleukins and interleukins receptors expression between CDC42 gene set statuses, consistent with previous study reports. Pegilodecakin (PEGylated recombinant IL_C_10) induces the proliferation of CD8 T cells both within the tumor microenvironment and in the systemic circulation, while also activating CD8 T cells within TME[51]. IL-21 functions as a robust survival factor for both natural killer (NK) and T cells, while also inhibiting the differentiation of Tregs[52]. Therefore, the significantly high expression of IL_C_10 and IL-21 in CDC42 gene set mutation patients may indicate the presence of more CD8 T cells in TME and a higher probability of a positive response to ICI therapy compared to patients with CDC42 gene set wild type status. Moreover, the association of IL-33 with the establishment of a tumorigenic niche has been reported[53], suggesting that its elevated expression in CDC42 wild type patients might imply a reduced likelihood of responding to ICI therapy. Overall, through the above analyses, we observed enhanced immunity and a greater probability response to ICI in CDC42 gene set mutation patients compared to CDC42 gene set wild type patients.

**Fig. 5.**
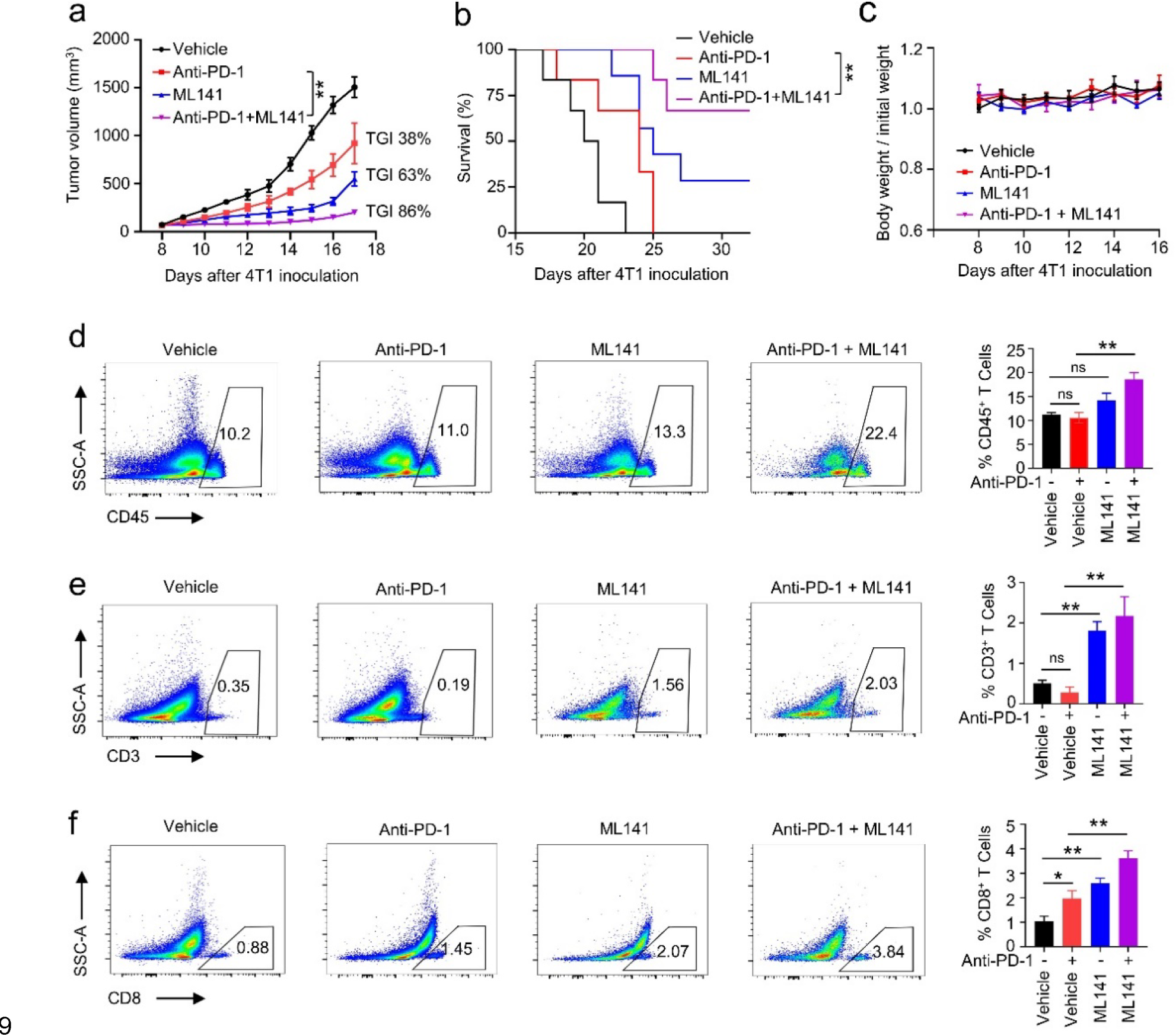
ML141 enhances Anti-PD-1 antibody induced tumor inhibition *in vivo*. **a**, Growth curves of tumors from the indicated groups (n = 8). **b**, Kaplan-Meier survival curves of mice in the indicated group (n = 8). **c**, Body weight change curves of mice in the indicated group. **d-f**, Impact of Anti-PD-1 antibody alone, ML141 alone, or a combination of both on the frequency of CD45^+^ lymphocytes, CD3^+^ T cells, and CD8^+^ T cells in TEM, assessed by flow cytometry. Error bars represent mean ± SEM; statistical analysis by Wilcoxon rank sum test (a) or logrank test (b) or two-tailed unpaired t-test (d-f). *, P < 0.05; **, P < 0.01; ns, no statistical difference, P > 0.05.

### Exploring the synergy of CDC42 inhibitor in combination with ICI to enhance immunotherapy efficacy

We investigated the antitumor effect of combining a CDC42 inhibitor (ML141) with anti-PD-1 antibody. Treatment with anti-PD-1 antibody alone demonstrated ordinary therapeutic capacity, with a TGI of 38% (Fig. 5a). Treatment with ML141 alone resulted in a TGI of 63% (Fig. 5a). Notably, the combination of ML141 and anti-PD-1 antibody significantly reduced tumor growth (TGI = 86%) and prolonged survival time compared to the anti-PD-1 antibody alone group (Fig. 5a and 5b). This suggests that the combination therapy can effectively exert potent antitumor immune activity. Furthermore, these treatments did not lead to weight loss in the mice (Fig. 5c), indicating that this dosing regimen is safe. To determine the role of the immune response in the antitumor activity of this dosage regimen, we analyzed the immune cells’ infiltration in the TME using flow cytometry experiments. The results showed that a combination of ML141 and an anti-PD-1 antibody significantly increased the frequency of CD45+ lymphocytes, CD3+ T cells, and CD8+ cytotoxic T cells in the TME compared to the anti-PD-1 antibody alone (Fig. 5d-f, Fig. S1). Furthermore, the use of ML141 alone also raised the frequency of CD3+ T cells and CD8+ cytotoxic T cells in the TME, aligning with earlier discoveries that the pharmacological inhibition of CDC42 triggers antitumor immune activity[4]. Interestingly, the CDC42 inhibitor mimics the defective function of CDC42 to a certain degree. As a result, our experiment confirms that the defective function of CDC42 is a biomarker for ICI therapy. Overall, these findings suggest that CDC42 inhibitor, used in conjunction with ICI, has a synergistic therapeutic effect and can enhance the efficacy of immunotherapy.

## Discussion

CDC42 downstream signals are known to be involved in stimulating tumors, including tumorigenesis, progression, invasion, and metastasis[3]. The purpose of this study is to investigate whether the defective function of CDC42 could be used as a biomarker for ICI therapy. We hypothesized that CDC42’s function may not only depend on CDC42 itself but also on its binding protein and effector protein. These genes are collectively referred to as the CDC42 gene set. Mutations in the CDC42 gene set genes may result in the partial defectiveness of CDC42 function. By analyzing a dataset of patients undergoing ICI treatment, we observed that patients with CDC42 gene set mutations had a higher rate of ORR and DCB. Additionally, these patients showed significantly longer OS and PFS time compared to patients with wild type CDC42 gene set genes. Bootstrap samples further confirmed that patients with CDC42 gene set mutations are more likely to respond to ICIs than patients with wild type CDC42 gene set genes. These analyses indicate that CDC42 gene set mutation can serve as a clinical biomarker for ICI therapy. In animal experiments, employing CDC42 inhibitors to mimic the defective function of CDC42 resulting in improved survival times in mice that were initially insensitive to anti-PD-1 treatment. This finding further validates the utility of the defective function of CDC42 as a potential biomarker for ICI treatment to some extent.

We conducted a further analysis of the CDC42 gene set’s status to better understand its role in indicating the clinical response to ICI using the TCGA dataset. Our findings revealed that tumors with CDC42 gene set mutations exhibited stronger immunogenicity, as evidenced by higher TMB and NAL. Additionally, we compared the gene expression levels of MHC1, TCR Shannon, and CYT, and observed significantly higher expression levels of these genes in patients with CDC42 gene set mutations. MHC1 plays a crucial role in presenting antigens to CD8 T cells, and its down-regulation has been associated with resistance to ICI[35]. Moreover, elevated level of TCR diversity and high expression of CYT indicate T cell activation and enhanced tumor cell killing efficiency. In summary, these comparisons highlight a more active immune response in CDC42 gene set mutation patients, characterized by stronger immunogenicity and the potential for more effective T cell activation and killing through increased MHC1 antigen presentation and TCR diversity, as well as higher CYT expression.

In TME, leukocyte fraction, lymphocyte fraction, TIL fraction and CD8 T cell levels were substantially higher in patients with CDC42 gene set mutations compared to those with CDC42 gene set wild type, indicating enhanced immunity. The increased expression of chemokines, such as CXCL9, CXCL10 and CXCL11, in patients with CDC42 gene set mutations can recruit more T cells into TME[47–49]. Additionally, higher expression of interleukins, like IL-10 and IL-21, in CDC42 gene set mutation patients promotes the survival of T cells. These findings suggest that CDC42 gene set mutation patients have a TME with increased infiltration of immune cell, particularly CD8 T cells, which experience improved survival conditions. Studies have shown that CD8 T cells play a critical role in eliminating tumors, and their presence within the tumor microenvironment is associated with better clinical responses to ICI treatments[34]. Furthermore, elevated expression of ICPs has been linked to positive responses to ICI treatments[33]. Therefore, the utilization of ICI therapies in patients with CDC42 gene set mutations (who exhibit elevated expression of ICPs) may potentially restore the suppressed function of CD8 T cells. In summary, the aforementioned analyses collectively highlight the significance of considering CDC42 gene set mutations as a potential biomarker for predicting responses to ICI treatment.

Some other biomarkers for ICI treatment, such as NOTCH4[54] and PAPPA2[55], do not indicate whether their biomarker function come from the alternation of their function, and the potential for combining their inhibitors with ICI was not examined. We hypothesize that the function of CDC42 may not only depend on itself but also its downstream binding protein and effector function. Mutations in CDC42 gene set result in the defective function of CDC42 and inhibition of tumor growth, which further releases immune suppression. Therefore, the functional defectiveness in CDC42 may underlie its potential as a biomarker for clinical benefit from ICI therapy. The significant difference between mutations in CDC42, its downstream binding proteins and effector proteins and the clinical benefit of ICI partially confirms our hypothesis. Hence, we propose that the combined use of ICI and CDC42 inhibitors could further enhance ICI’s efficacy, and we conducted experiment exploration. Animal experiment data demonstrated that the ML141 inhibitor indeed could further promote ICI’s efficacy, further confirming our hypothesis and providing valuable insights for further exploration in clinical settings.

## Conclusion

In conclusion, we have speculated that CDC42’s function may depend not only on CDC42 itself but also its binding proteins and effector proteins. We have also demonstrated that mutations in the CDC42 gene set could serve as a novel biomarker for predicting the clinical response of ICI therapy. Furthermore, the analysis of the TCGA dataset and the animal experiment further supports the role of this predictive biomarker. Additionally, this study has provided insight into the potential synergistic effects of combining CDC42 inhibitors with ICIs to enhance their efficacy, especially bringing hope to situations that failed to respond to anti-PD-1 treatment. Our study introduces a novel approach to biomarker analysis, considering that changes in the function of a key gene can result from mutations in downstream effector proteins as well. Analyzing gene sets collectively like this may facilitate the discovery of new biomarkers and potential drug targets.

## Data Availability

All data produced in the present work are contained in the manuscript

## Abbreviations

CI: confidence interval
CR: complete response
CTLA-4: cytotoxic T lymphocyte-associated protein-4
CYT: cytolytic activity
DCB: Durable clinical benefit
GSEA: gene set enrichment analysis
GZMA: granzyme A
HCC: hepatocellular carcinoma
HR: hazard ratio
ICIs: immune checkpoint inhibitors
ICP: immune checkpoint
KEGG: Kyoto Encyclopedia of Genes and Genomes
KM: Kaplan-Meier
NAL: neoantigen load
NE: non-evaluable
NR: non-responder
ORR: objective response rate
OS: overall survival
PD: progressive disease
PD-1: Programmed Death Receptor 1
PFS: progression-free survival
PFS: progression-free survival
PR: partial response
PRF1: perforin 1
R: responder
Rho: ras homologous
SD: stable disease
SNV: single nucleotide variation
ssGSEA: single-sample gene set enrichment analysis
STD: standard deviation
TCGA: The Cancer Genome Atlas
TCGA: The Cancer Genome Atlas
TIL: tumor-infiltrating lymphocyte
TMB: tumor mutational burden
TME: tumor microenvironment
Tregs: regulatory T cells
VEGF: vascular endothelial growth factor
WES: whole exome sequencing

## Declarations

### Ethics approval and consent to participate

Not applicable.

### Consent for publication

Not applicable.

### Availability of data and materials

The datasets supporting the conclusions of this article are included within the article.

### Competing interests

The authors declare that they have no competing interests.

### Funding

We gratefully acknowledge financial support from National Natural Science Foundation of China (T2225002, 82273855 to M.Y.Z.), SIMM-SHUTCM Traditional Chinese Medicine Innovation Joint Research Program (E2G805H), Shanghai Municipal Science and Technology Major Project, National Key Research and Development Program of China (2022YFC3400504 to M.Y.Z.), the Youth Innovation Promotion Association CAS (2023296 to S.L.Z.) and the Natural Science Foundation of Shanghai (22ZR1474300 to S.L.Z.).

### Authors contributions

M.Y.Z, S.L.Z, K.W and Y.Y. Zhang conceived of and designed the study. K.W and Y.Y. Zhang drafted the manuscript. K.W conducted the data analysis. Y.Y. Zhang and Z.M.S performed the experiments. C.Y.X provided administrative support. B.W, Y.Y. Zhou, X.C.T and X.M.L participated in the discussion. All authors read and approved the final version of the manuscript and are accountable for all aspects of the work.

## Acknowledgements

The authors would like to acknowledge their colleague, mentor, and friend, Dr. Hualiang Jiang (1965–2022), who took part in the work and in the preparation of the original manuscript.

## Notes

### Competing Interest Statement

The authors have declared no competing interest.

### Author Declarations

Immunotherapy treatment datasets were obtained from https://www.cbioportal.org and https://www.sciencedirect.com/science/article/pii/S0092867417311224?via%3DihubTCGA datasets were obtained from https://portal.gdc.cancer.gov/ and https://www.sciencedirect.com/science/article/pii/S0092867418302290#mmc1

## Reference

1. Sharma, P., et al., Primary, Adaptive, and Acquired Resistance to Cancer Immunotherapy. Cell, 2017. 168(4): p. 707–723.

2. Cancer immunotherapy: the quest for better biomarkers. Nat Med, 2022. 28(12): p. 2437.

3. Xiao, X.H., et al., Regulating Cdc42 and Its Signaling Pathways in Cancer: Small Molecules and MicroRNA as New Treatment Candidates. Molecules, 2018. 23(4).

4. Kalim, K.W., et al., Targeting of Cdc42 GTPase in regulatory T cells unleashes antitumor T-cell immunity. J Immunother Cancer, 2022. 10(11).

5. Xu, J., et al., Serum cell division cycle 42 in advanced hepatocellular carcinoma patients: Linkage with clinical characteristics and immune checkpoint inhibitor-related treatment outcomes. Clin Res Hepatol Gastroenterol, 2023. 47(7): p. 102149.

6. Guo, L., et al., Serum cell division cycle 42 reflects the treatment response and survival in patients with advanced cervical cancer who receive immune checkpoint inhibitor treatment. Oncol Lett, 2023. 26(3): p. 414.

7. De Henau, O., et al., Overcoming resistance to checkpoint blockade therapy by targeting PI3Kgamma in myeloid cells. Nature, 2016. 539(7629): p. 443-447.

8. Chen, H.Y., et al., Inhibition of redox/Fyn/c-Cbl pathway function by Cdc42 controls tumour initiation capacity and tamoxifen sensitivity in basal-like breast cancer cells. EMBO Mol Med, 2013. 5(5): p. 723–36.

9. Miao, D., et al., Genomic correlates of response to immune checkpoint therapies in clear cell renal cell carcinoma. Science, 2018. 359(6377): p. 801-806.

10. Hugo, W., et al., Genomic and Transcriptomic Features of Response to Anti-PD-1 Therapy in Metastatic Melanoma. Cell, 2016. 165(1): p. 35–44.

11. Riaz, N., et al., Tumor and Microenvironment Evolution during Immunotherapy with Nivolumab. Cell, 2017. 171(4): p. 934–949 e16.

12. Miao, D., et al., Genomic correlates of response to immune checkpoint blockade in microsatellite-stable solid tumors. Nat Genet, 2018. 50(9): p. 1271–1281.

13. Rizvi, N.A., et al., Cancer immunology. Mutational landscape determines sensitivity to PD-1 blockade in non-small cell lung cancer. Science, 2015. 348(6230): p. 124-8.

14. Snyder, A., et al., Genetic basis for clinical response to CTLA-4 blockade in melanoma. N Engl J Med, 2014. 371(23): p. 2189–2199.

15. Van Allen, E.M., et al., Genomic correlates of response to CTLA-4 blockade in metastatic melanoma. Science, 2015. 350(6257): p. 207–211.

16. Hellmann, M.D., et al., Genomic Features of Response to Combination Immunotherapy in Patients with Advanced Non-Small-Cell Lung Cancer. Cancer Cell, 2018. 33(5): p. 843–852 e4.

17. Liu, D., et al., Integrative molecular and clinical modeling of clinical outcomes to PD1 blockade in patients with metastatic melanoma. Nat Med, 2019. 25(12): p. 1916–1927.

18. Zhang, Z., et al., EPHA7 mutation as a predictive biomarker for immune checkpoint inhibitors in multiple cancers. BMC Med, 2021. 19(1): p. 26.

19. Colaprico, A., et al., TCGAbiolinks: an R/Bioconductor package for integrative analysis of TCGA data. Nucleic Acids Res, 2016. 44(8): p. e71.

20. Liu, J., et al., An Integrated TCGA Pan-Cancer Clinical Data Resource to Drive High-Quality Survival Outcome Analytics. Cell, 2018. 173(2): p. 400–416 e11.

21. Thorsson, V., et al., The Immune Landscape of Cancer. Immunity, 2018. 48(4): p. 812–830 e14.

22. Villaruz, L.C. and M.A. Socinski, The clinical viewpoint: definitions, limitations of RECIST, practical considerations of measurement. Clin Cancer Res, 2013. 19(10): p. 2629–36.

23. Rizvi, H., et al., Molecular Determinants of Response to Anti-Programmed Cell Death (PD)-1 and Anti-Programmed Death-Ligand 1 (PD-L1) Blockade in Patients With Non-Small-Cell Lung Cancer Profiled With Targeted Next-Generation Sequencing. J Clin Oncol, 2018. 36(7): p. 633–641.

24. Saltz, J., et al., Spatial Organization and Molecular Correlation of Tumor-Infiltrating Lymphocytes Using Deep Learning on Pathology Images. Cell Rep, 2018. 23(1): p. 181–193 e7.

25. He, Y., et al., Classification of triple-negative breast cancers based on Immunogenomic profiling. J Exp Clin Cancer Res, 2018. 37(1): p. 327.

26. Hänzelmann, S., R. Castelo, and J. Guinney, GSVA: gene set variation analysis for microarray and RNA-seq data. BMC bioinformatics, 2013. 14: p. 1–15.

27. Love, M.I., W. Huber, and S. Anders, Moderated estimation of fold change and dispersion for RNA-seq data with DESeq2. Genome Biol, 2014. 15(12): p. 550.

28. Yu, G., et al., clusterProfiler: an R package for comparing biological themes among gene clusters. OMICS, 2012. 16(5): p. 284–7.

29. Rooney, M.S., et al., Molecular and genetic properties of tumors associated with local immune cytolytic activity. Cell, 2015. 160(1-2): p. 48–61.

30. Ruiz-Bañobre, J., et al., Rethinking prognostic factors in locally advanced or metastatic urothelial carcinoma in the immune checkpoint blockade era: a multicenter retrospective study. ESMO open, 2021. 6(2): p. 100090.

31. Bland, J.M. and D.G. Altman, The logrank test. Bmj, 2004. 328(7447): p. 1073.

32. Virtanen, P., et al., SciPy 1.0: fundamental algorithms for scientific computing in Python. Nat Methods, 2020. 17(3): p. 261–272.

33. Hu, F.F., et al., Expression profile of immune checkpoint genes and their roles in predicting immunotherapy response. Brief Bioinform, 2021. 22(3).

34. Oba, T., et al., Overcoming primary and acquired resistance to anti-PD-L1 therapy by induction and activation of tumor-residing cDC1s. Nat Commun, 2020. 11(1): p. 5415.

35. Luo, N., et al., DNA methyltransferase inhibition upregulates MHC-I to potentiate cytotoxic T lymphocyte responses in breast cancer. Nat Commun, 2018. 9(1): p. 248.

36. Johnson, D.B., et al., Melanoma-specific MHC-II expression represents a tumour-autonomous phenotype and predicts response to anti-PD-1/PD-L1 therapy. Nat Commun, 2016. 7: p. 10582.

37. Chen, L. and D.B. Flies, Molecular mechanisms of T cell co-stimulation and co-inhibition. Nature Reviews Immunology, 2013. 13(4): p. 227–242.

38. Jeong, S. and S.H. Park, Co-Stimulatory Receptors in Cancers and Their Implications for Cancer Immunotherapy. Immune Netw, 2020. 20(1): p. e3.

39. Liu, R., et al., Influence of Tumor Immune Infiltration on Immune Checkpoint Inhibitor Therapeutic Efficacy: A Computational Retrospective Study. Front Immunol, 2021. 12: p. 685370.

40. Li, B., Why do tumor-infiltrating lymphocytes have variable efficacy in the treatment of solid tumors? Front Immunol, 2022. 13: p. 973881.

41. Zhang, H., et al., Roles of tumor-associated macrophages in anti-PD-1/PD-L1 immunotherapy for solid cancers. Mol Cancer, 2023. 22(1): p. 58.

42. Jiang, M., et al., Alterations of DNA damage response pathway: Biomarker and therapeutic strategy for cancer immunotherapy. Acta Pharm Sin B, 2021. 11(10): p. 2983–2994.

43. Ward, A.B., et al., Enhancing anticancer activity of checkpoint immunotherapy by targeting RAS. MedComm (2020), 2020. 1(2): p. 121-128.

44. Li, X., et al., Navigating metabolic pathways to enhance antitumour immunity and immunotherapy. Nature Reviews Clinical Oncology, 2019. 16(7): p. 425–441.

45. Ma, W., B. Pham, and T. Li, Cancer neoantigens as potential targets for immunotherapy. Clin Exp Metastasis, 2022. 39(1): p. 51–60.

46. Johnson, B.J., et al., Single-cell perforin and granzyme expression reveals the anatomical localization of effector CD8+ T cells in influenza virus-infected mice. Proceedings of the National Academy of Sciences, 2003. 100(5): p. 2657–2662.

47. Peng, D., et al., Epigenetic silencing of TH1-type chemokines shapes tumour immunity and immunotherapy. Nature, 2015. 527(7577): p. 249-53.

48. Reschke, R. and T.F. Gajewski, CXCL9 and CXCL10 bring the heat to tumors. Science immunology, 2022. 7(73): p. eabq6509.

49. Li, Y., et al., CXCL11 Correlates with Immune Infiltration and Impacts Patient Immunotherapy Efficacy: A Pan-Cancer Analysis. Front Immunol, 2022. 13: p. 951247.

50. Hsieh, C.H., et al., Potential Role of CXCL13/CXCR5 Signaling in Immune Checkpoint Inhibitor Treatment in Cancer. Cancers (Basel), 2022. 14(2).

51. Tannir, N.M., et al., Pegilodecakin as monotherapy or in combination with anti-PD-1 or tyrosine kinase inhibitor in heavily pretreated patients with advanced renal cell carcinoma: Final results of cohorts A, G, H and I of IVY Phase I study. Int J Cancer, 2021. 149(2): p. 403–408.

52. Mortezaee, K. and J. Majidpoor, Checkpoint inhibitor/interleukin-based combination therapy of cancer. Cancer Med, 2022. 11(15): p. 2934–2943.

53. Briukhovetska, D., et al., Interleukins in cancer: from biology to therapy. Nat Rev Cancer, 2021. 21(8): p. 481–499.

54. Long, J., et al., Identification of NOTCH4 mutation as a response biomarker for immune checkpoint inhibitor therapy. BMC Med, 2021. 19(1): p. 154.

55. Dong, Y., et al., PAPPA2 mutation as a novel indicator stratifying beneficiaries of immune checkpoint inhibitors in skin cutaneous melanoma and non-small cell lung cancer. Cell Prolif, 2022: p. e13283.

