## Supplementary Material for "Mutation in CDC42 gene set as a response biomarker for immune checkpoint inhibitor therapy"

| **Data sets** | **Target** | **Cancer type** | **Patient number** |
| --- | --- | --- | --- |
| Miao2019 cohort | Anti-PD-1 | Renal Clear Cell Carcinoma | 35 |
| Hugo cohort | Anti-PD-1 | Melanoma | 38 |
| Miao2018 cohort | Anti-CTLA-4  Anti-PD-1  Anti-CTLA-4 + Anti-PD-1 | Non-Small Cell Lung Cancer  Bladder Cancer  Melanoma  Head and Neck Cancer | 249 |
| Rizvi cohort | Anti-PD-1 | Non-Small Cell Lung Cancer | 35 |
| Snyder cohort | Anti-CTLA-4 | Melanoma | 64 |
| Van Allen cohort | Anti-CTLA-4 | Melanoma | 110 |
| Riaz cohort | Anti-PD-1 | Melanoma | 73 |
| Hellmann cohort | Anti-CTLA-4 + Anti-PD-1 | Non-Small Cell Lung Cancer | 75 |
| Liu cohort | Anti-PD-1 | Melanoma | 144 |

**Table S1. Description of WES Data sets that received ICI therapy.**

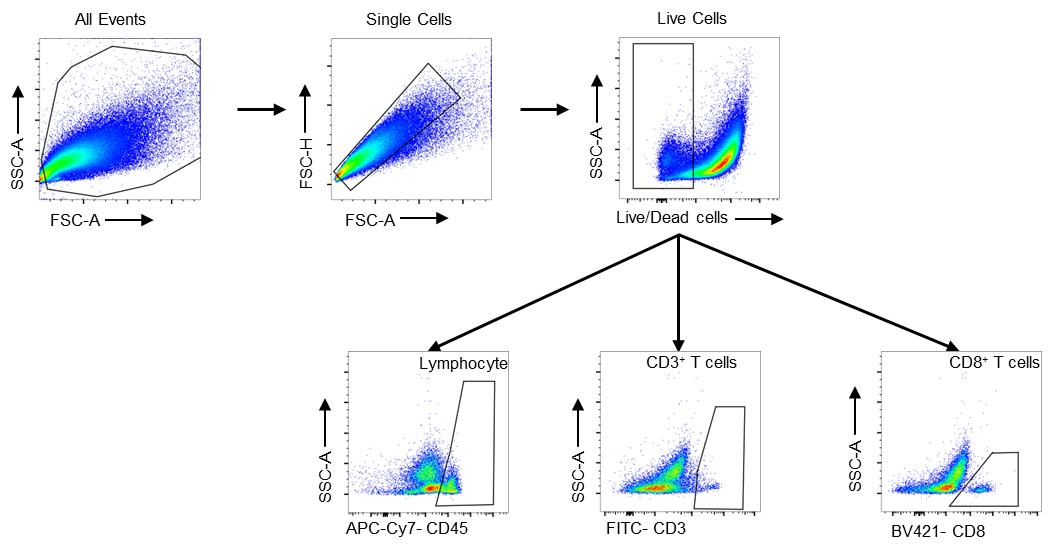

**Figure S1.** Flow cytometry gating strategies.
